# Transient increased risk of influenza infection following RSV infection in South Africa; findings from the PHIRST study, South Africa, 2016-2018

**DOI:** 10.1101/2023.05.30.23290741

**Authors:** Naomi R Waterlow, Jackie Kleynhans, Nicole Wolter, Stefano Tempia, Rosalind M Eggo, Orienka Hellferscee, Limakatso Lebina, Neil Martinson, Ryan G Wagner, Jocelyn Moyes, Anne von Gottberg, Cheryl Cohen, Stefan Flasche

## Abstract

Large-scale prevention of respiratory syncytial virus (RSV) infection may have ecological consequences for co-circulating pathogens, including influenza. We assessed if and for how long RSV infection alters the risk for subsequent influenza infection.

We analysed a prospective longitudinal cohort study conducted in South Africa between 2016 and 2018. For participating households, nasopharyngeal samples were taken twice weekly, irrespective of symptoms, across three respiratory virus seasons, and real-time polymerase chain reaction (PCR) was used to identify infection with RSV and/or influenza. We fitted an individual-level hidden markov transmission model in order to estimate RSV and influenza infection rates and their interdependence.

Of a total of 122113 samples collected, 1265 (1.0%) were positive for influenza and 1002 (0.8%) positive for RSV, with 15 (0.01%) samples from 12 individuals positive for both influenza and RSV. We observed 2.25-fold higher incidence of co-infection than expected if assuming infections were unrelated. We estimated that infection with influenza is 2.13 (95% CI 0.97 - 4.69) times more likely when already infected with, and for a week following, RSV infection, adjusted for age. This equates to 1.4% of influenza infections that may be attributable to RSV in this population. Due to the local seasonality (RSV season precedes the influenza season), we were unable to estimate changes in RSV infection risk following influenza infection.

RSV infection was associated with an increased risk for influenza infection for a short period after infection. However, the impact on population-level transmission dynamics of this individual-level synergistic effect was not measurable in this setting.

**Research in Context:** *Evidence before this study:* We searched PubMed titles and abstracts for the terms “influenza”, “RSV” or “Respiratory syncytial virus” and “interaction”, “competition” or “enhancement” resulting in 56 articles, excluding reviews. Evidence for the potential interaction of influenza and RSV originates from analyses of viral surveillance and experimental non-human studies, or isolated mathematical models. Most such studies suggest potential competitive exclusion of RSV and Influenza but are prone to potential confounding and unable to test the links between biological mechanisms and population level impacts.

*Added value of this study:* This longitudinal study with frequent testing of participants for colonisation with RSV and Influenza allows sufficient resolution to analyse direct evidence for interaction of both viruses on colonisation. In contrast to evidence insofar we find that the effect of RSV colonisation on the risk for influenza acquisition is short lived and synergistic, but unlikely to substantially effect influenza epidemiology on population level.

*Implications of all the available evidence:* RSV infections are likely to have limited impact on influenza circulation.

## Background

Globally, in 2019 there were an estimated 17.2 billion upper respiratory tract infections (1), which is a common presentation of the over 200 known respiratory virus strains that cause illness in humans(1). These viruses may interact, resulting in cross-protective or enhancing effects for transmission and/or disease severity(2–5), and can result in unintended knock-on effects from public health interventions. However, other factors, including a change in behaviour relevant to transmission, may also underlie apparent pathogen interaction. An example of ecological knock-on effects that were not caused by pathogen interaction but by a change in social behaviour has been observed as a result of social distancing to mitigate the Coronavirus Disease 2019 (COVID-19) pandemic(6). Vaccination may increase the burden of an untargeted virus if an infection with the targeted pathogen is cross-protective, or reduce the prevalence of enhancing pathogens. Understanding these potential ecological knock-on effects is particularly important for pathogens where vaccines are in late-stage clinical development, such as for respiratory syncytial virus (RSV) where successful Phase III trials have recently been reported for multiple vaccines(7,8).

While RSV may be a critical factor underlying a substantial part of severe pneumococcal infections(9), some evidence has suggested a competition for a similar ecological niche with influenza viruses(5). However, much of the available evidence is based on *in vitro* experiments or population-level ecological studies that identify the correlation of respective case series; e.g. following the 2009 influenza pandemic, many studies showed a delayed or absent RSV epidemic(10–14), however, this was not uniformly the case(15). Surveillance in non-pandemic years has shown that the epidemic incidence peaks of influenza and RSV rarely coincide in geographies where influenza and RSV circulate at the same time of year (16,17). Immunological evidence from studies such as cell culture and mouse models suggest potential inhibition of heterologous growth(18–21), but the extent that such interactions impact transmission among humans is unclear.

A key issue faced by many epidemiological studies of RSV and influenza interaction is that they rely on syndromic surveillance of medically attended illness for their inference, even though asymptomatic infections and non-medically attended illness are likely important drivers of transmission and interaction. We analyse a unique longitudinal household study with frequent asymptomatic sampling to help better understand the interdependence of RSV and influenza infection.

## Methods

### Study population and data collection

Influenza and RSV infections were identified as part of the Prospective Household cohort study of influenza, respiratory syncytial virus and other respiratory pathogens community burden and Transmission dynamics in South Africa (PHIRST) described in detail elsewhere (22). The study was conducted between 2016 and 2018 in both a rural and an urban community in South Africa. Households of more than two individuals were enrolled if 80% or more individuals from the household provided consent. The study was conducted over three respiratory virus seasons, with new households enrolled each year for a follow-up period of ten months, except in the 2016 season, where only 6 months of follow-up occurred, starting in the middle of the RSV season (Figure 1). Nasopharyngeal swabs were collected twice per week, irrespective of symptoms, and samples were subsequently analysed by real-time polymerase chain reaction (PCR) for the presence of RSV and influenza viruses.

**Figure 1:**
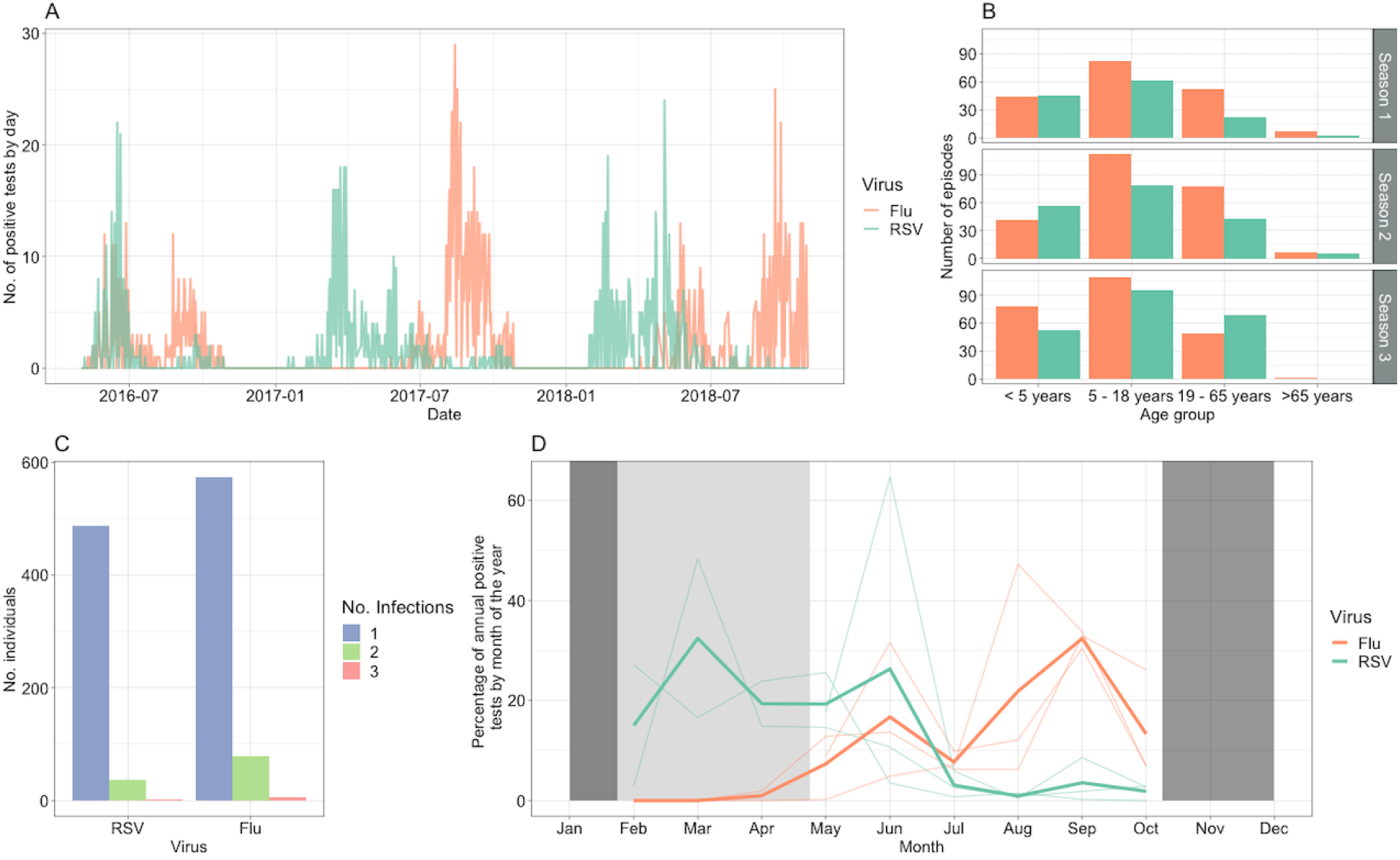
A) All positive test results over time for influenza (Flu) and Respiratory Syncytial Virus (RSV). B) Number of infection episodes in each age group, across the three seasons C) Number of individuals against the number of infections for each virus, where positive tests within 14 days are assumed to be due to the same infection. D) Seasonality of influenza and RSV infections, depicting the percentage of annual positive tests by month of the year. The semi-transparent lines depict the individual seasons. The solid lines depict the mean across seasons. Dark shaded areas depict months where no samples were taken, and the light shaded area shows samples not taken in the 2016 season.

We defined an ‘episode’ as one or more positive samples that do not have more than 14 days of negative samples between them (23,24). We define ‘dual infection’ as being infected with both viruses at the same time (i.e. the same sample is positive for both RSV and influenza). We defined ‘overlapping episodes’ as when an RSV episode coincided at some point in time with an influenza episode.

### Crude Analyses

For the crude analysis, we included only the first positive sample for each episode, so as not to double-count infections. For each individual with an RSV infection (i.e. episode) in the dataset, we identified all controls who were tested but uninfected on the same day and were in the same age group (defined as ages <5, 5 to 18, 19-65 and >65 years). We then calculated the number of individuals among cases (exposed) and controls (unexposed) that had a positive sample for influenza infection (outcome variable) on the day of the first positive RSV sample or up to 21 days before or after. This range was used as we consider it a biologically plausible range for interaction. We repeated this process in reverse, identifying RSV-positive samples on the same and nearby days as an influenza episode started. Risk ratios and the respective confidence intervals were then calculated across weekly aggregated counts (see supplement).

Secondly, we compared the observed frequency of samples indicating RSV and influenza co-infection versus the expected frequency of co-infection if assuming that the risk of RSV and influenza infection were uncorrelated. The expected proportion of co-infections on a given day is calculated by multiplying the prevalence of RSV and influenza infections on that day. The expected proportion of co-infections overall is then the sum of expected co-infection frequency on the sampling days (proportion times the number of samples) divided by the total number of samples taken during the study. We compared the expected proportion of co-infected samples against the observed proportion of co-infected samples using a two-proportion z-test. All analysis was conducted in R.

### Model

We used a multi-state Markov modelling framework to model the transition between RSV, influenza and co-infection states (25). Each individual is classed as either Susceptible (S), Infectious (I), Period of interaction following infectiousness (P) or Recovered (R) (Figure 2). We fit parameters for the time-dependent Force of Infection (FOI) of both viruses and the strength of interaction for each virus. The time dependence of the FOI aims to account for the timing of the relative RSV and influenza seasons, assuming this may be related to some unmeasured factors. Interaction can occur during, and for one week after infection, as this was the scale of time indicated in previous studies (26). We also ran sensitivity analyses with 2, 3 and 4 weeks, respectively. The interaction can be competitive (<1 indicating that infection with one partially mitigates acquisition of the other) or synergistic (>1 indicating that infection with one enhances the probability of acquisition with the other pathogen), and we test the null hypothesis of there being no interaction. Other parameters were informed from literature estimates (Table 1). The FOI is time-dependent, independently estimated every 30-day time window, and age group is included as a covariate (<5 years, 5 -18 years, 19-65 years, 65 years or older). Further details are given in the supplement.

**Figure 2:**
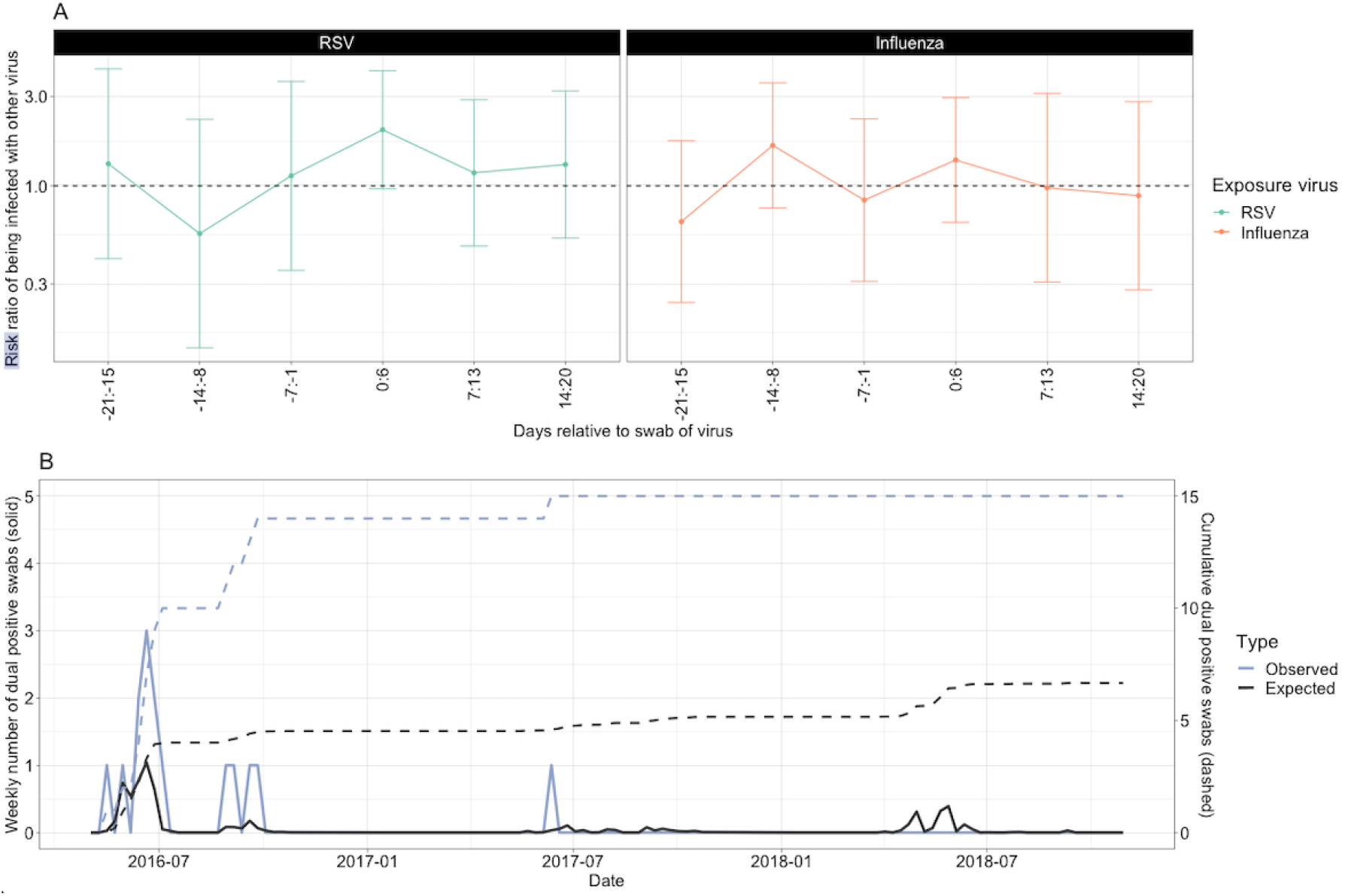
A) Analysis 1. Risk ratio of the risk of being infected by the second virus (outcome), when exposed to the primary virus. Points indicated the estimate and error bars the 95% confidence interval. The dashed line indicates a risk ratio of 0, equating to no effect. B) Analysis 2. Expected versus observed dual infections per week.

We assumed that all individuals were susceptible to both RSV and influenza infection at recruitment (S_RSV_S_INF_ class in the model), and that each individual could only be infected by each virus once during a season. If a further infection occurred, data on the individual was removed from the time point of the re-infection. We assumed that positive tests of the same virus occurring within a 14-day time window were related to the same infection(27).

The observation of a current RSV or influenza infection in the study did not allow us to distinguish the period of post-infection interaction from the subsequent state of immunity without interaction, so in the model, these were included as “censored” states. This means the model estimates which of these states each individual is in at a given time.

In addition, we assumed that all RSV and influenza positive tests were true positives, while we estimated the proportion of false negative tests in the model framework. We did this by allowing misclassification of states for negative samples.

The model was implemented and fit using the R *msm* package(25), with the quasi-Newton method “*BFGS”*, which builds up a picture of the surface to be optimised using function values and gradients. We ran the model twice, with different initial values each time.

### Sensitivity Analysis

We tested the sensitivity of our results to alternative assumptions of the duration of interaction, rerunning the model where interaction can occur during infection and for the following 2 weeks, the following 3 weeks and the following 4 weeks. We also attempted to run the model with shorter time intervals for the FOI, as well as excluding the first season (2016) where most of the co-infections occurred, however neither of these models reached convergence.

All analysis was conducted in R. Code and summarised data are available here https://github.com/NaomiWaterlow/markov_model.

## Results

Out of a total number of 122113 individual-days swab collection was set to occur (across 1684 individuals), 13% of results were missing from the analysis, either due to unavailability of the individuals or processing errors (such as incorrect labelling or leakage). In total, there were 1265 positive influenza samples and 1002 positive RSV samples, with a median of 71 samples taken per individual (95% quantiles 25-81) over the study period. These clustered into 574 and 488 episodes of infection with influenza or RSV, of which 14% (83/574) and 8% (40/488), respectively, were episodes of reinfection of an individual with the same pathogen within a season (Figure 1 A&C). There were 15 dual infection samples across 12 episodes of infection, in 12 individuals: two in children <5 years old, nine in children 5-18 years old and one in an adult 19-65 years old. There were 18 samples that were classified as overlapping, corresponding to the same 12 infections and individuals as the dual infections, with no further overlapping episodes that were not detected as a dual infection.

The RSV epidemics occurred from February to June, with peak incidence around March, whilst the influenza epidemic occurred later in the year between May and October, typically peaking in September (Figure 1D). The mean duration of episodes (from first to last positive samples) was 6.8 days for RSV (95% CI 6.1 - 7.4 days) and 6.5 days for influenza (95% CI 6.0 - 7.0 days). The infection attack rate for each season was 34% (216/512), 41% (216/577) and 42% (216/565) for influenza and 24% (131/542), 32% (184/577) and 38% (216/565) for RSV, although data collection over the first season was for a shorter period, missing much of the RSV season. The median age of infected individuals for influenza and RSV in the first season was 10 and 7 years, respectively (ranges 0-79 and 0-70), compared to 15 and 19 years in the second season (ranges 0-91 and 0-91) and 8 and 11 years in the third season (ranges 0-70 and 0-91) (Figure 1B).

In the crude analysis, a small increase in the risk ratio for influenza infection was observed the week following a positive RSV sample (1.99, 95%CI 0.96 - 4.11) (Figure 2A), although confidence intervals crossed one. Whilst there was also an increased risk ratio for RSV infection in the week following influenza infection, this was of a smaller magnitude and had lower confidence (1.37 95%CI 0.64 - 2.95).

Assuming independence of RSV and influenza infection, we would expect 6.67 co-infected samples to occur during the study period, compared to 15 that were observed; a 2.25 times higher rate of co-infections albeit not statistically significant (95%CI 0.73 - 3.77, p-value 0.12). All but one of the actual co-infections occurred in the first season (Figure 2B).

As the RSV season preceded the influenza season, only 15 /1684 (0.9%) study participants were infected with influenza and a subsequent RSV infection in the same season. Thus, in the Markov model, the estimated strength of interaction exerted by infection of influenza on RSV infection probability had wide confidence intervals, which crossed 1 (the Null hypothesis indicating no effect): RSV infection was 0.93 (95% CI: 0.2 - 4.3) times as likely in the presence of influenza infection compared to without influenza infection.

Conversely, 158/1684 (9.4%) individuals were infected with RSV and, subsequently, influenza. We estimated a synergistic interaction effect of RSV infection on the risk for influenza infection: infection with influenza was 2.13 (95% CI: 0.97 - 4.69) times more likely for one week following the first positive sample of an RSV infection episode. This translates to a very small population-level effect: of the 574 observed influenza episodes, only 12 were overlapping infections. If we add to these the infections that occurred within 7 days of testing negative after an infection episode with the other virus, we get a total of 18 extended overlap episodes. Eight of these 18 overlapping episodes would have been avoided without the synergistic effect, resulting in a reduction in total influenza cases in this cohort of 1.4%.

The model further estimated that the sensitivity of testing for influenza and RSV was very high, with a false negative rate of 0.75% (95% CI: 0.72 - 0.77). For RSV, those <5 years had the highest risk of being infected, followed by individuals aged 5-18 years (hazard ratio 0.51 (95% CI: 0.41- 0.63)), 19-65 years (hazard ratio 0.29 (95% CI: 0.23- 0.37)) and 65 years and older (hazard ratio 0.13 (95% CI: 0.06-0.30)) (Figure 3B). A similar but smaller reduction in infection risk with increasing age was observed for influenza (Figure 4).

**Figure 3:**
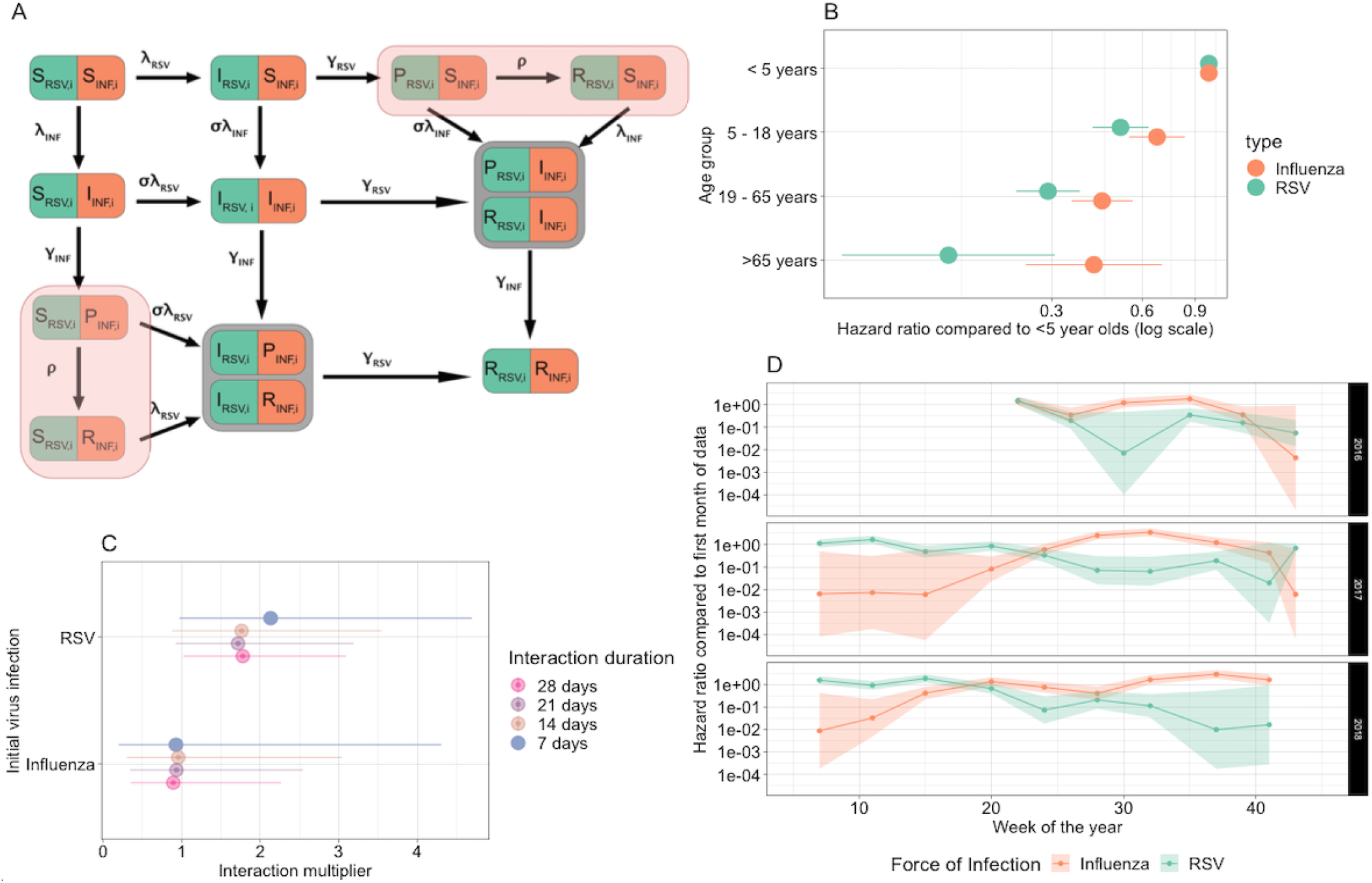
A) Model diagram. Shaded red compartments are censored states. B) Age group hazard ratios of infection by age on a log scale, pared to the <5 years as the base age group. C) Interaction multiplier for each virus, for the main analysis (7 days interaction), and the duration sensitivity analysis. An interaction multiplier of x indicates that Transmission from S to I with the other virus is x times more likely when already infected with the initial virus. D) Force infection over time for influenza and RSV (7-day interaction model). Lines indicate the mean value, and the shaded ribbons the 95% CI.

Our results on the strength of the synergistic effect of RSV infection on the risk for influenza infection were robust to alternative assumptions on the duration of said effect. We estimated that influenza infection is 1.76 (95%CI 0.88 - 3.54) times, 1.72 (95%CI 0.92 - 3.19) times and 1.78 (1.02 - 3.09) times more likely following an RSV infection if we assumed that the interaction lasted for 2, 3 or 4 weeks respectively (See supplement).

Our sensitivity analyses with a shorter time interval for the changing FOI, and for the default model but excluding the first season of data did not converge.

## Discussion

We found over twice as many co-infections than expected, if assuming no interaction, and similarly estimate in the model that influenza infection risk is 2.13 (95% CI: 0.97 - 4.69) higher during or shortly after RSV infection. This, however, translates to a very small population-level impact. Whilst we did not find evidence to suggest elevated RSV infection risk following influenza infection, this may have been due to the small sample size of RSV infections subsequent to influenza. Since the observed synergy in infection risk was estimated to be short-lived, ecological consequences on influenza infection risk following widespread RSV prevention are unlikely; however, they may be amplified by increases in severity as a result of co-infection(2), which we did not address in this paper.

The major strength of this analysis is that the data consist of regular symptom-agnostic swabbing of study participants, allowing the inclusion of mild and asymptomatic infections (which can be a large proportion), rather than just symptomatic and severe cases. This high-quality data results in major benefits. Firstly, changes in infection risk may be obscured by the impacts of dual infection on clinical severity when using syndromic surveillance data, resulting in higher reporting. This has been avoided in our study. The current literature gives mixed results on the increased severity of dual infections, with some studies estimating an increased severity of dual infections (26,28), and others not finding evidence of a significant effect (29,30). The impact of RSV and influenza co-infection on the severity of infection was outside the scope of this study.

A second benefit of symptom-agnostic swabbing is that no bias is introduced as a result of the inherent difference between those with symptoms and those without. For instance, it could be that those with more severe symptoms may have weaker immune systems than those with milder symptoms, and therefore may also be more susceptible to infection with both viruses. Including all infections in the analysis removes this potential bias.

Thirdly, by fitting models to infections rather than clinical cases, we reduce the number of parameters required to be fitted, as we do not need to include reporting rate parameters. Fitting complex interaction models is difficult, and it is not possible to differentiate extreme interaction parameter values from less extreme ones(31).

A further strength of the data used in this model is its longitudinal nature. We used data across almost 3 seasons, where the same individuals were sampled within (but not across) each season, giving us detailed infection data. There may, however, be bias introduced due to missing data, with 13% of swabs missing. Whilst the majority of these are likely due to unbiased reasons (such as travel and holidays), other potential reasons, such as severe infections resulting in hospitalisations, may have resulted in bias. However, due to the regularity of the swabbing, any bias introduced due to these reasons would have been minimal, as infection would likely have been identified on a neighbouring swab date (as the median recorded episode duration was 6 and 5.5 days for RSV and influenza, respectively) and very few hospitalisations were found in the cohorts overall. In addition, the hidden Markov model inference takes into account missing data.

Our model results differ from some previous evidence of interaction between RSV and influenza. However, these earlier findings are not based on mechanistic models and could therefore be confounded by other transmission-relevant factors, such as contact rates. This is likely the case with studies investigating causality for the shifts in RSV patterns following the 2009 influenza pandemic (10,12–14), where, despite no governmentally imposed social restrictions, fear of the virus could have altered contact behaviours (32). In our previous work on evidence for interaction between the two viruses in Nha Trang, Vietnam, we showed that the data was compatible with either a ∼41% reduction in susceptibility for 10 days following infection with the other virus, or no reduction in susceptibility following infection(26). This is not incompatible with our current estimate of a small increase in susceptibility on the individual level, which would likely not have an impact on the population scale. Our previous model used population-level symptomatic surveillance data, and we did not account for the possibility of cross-protective interaction, as opposed to this study, where the individual-level data allowed us to explore a broader parameter space.

Age-related susceptibility to infection reduction is generally considered to be the case for RSV, whether this be due to age directly, or due to subsequent infections. For instance, Henderson et al. showed that attack rates during an epidemic were 98% for the 1st infection, 75% for a second infection and 65% for a third infection (33). As increasing numbers of previous infections will strongly correlate with age, this is not out of line with our estimates of age-related hazard rates.

Whilst the evidence we provide shows an increased likelihood of influenza infection following RSV, this is of very small magnitude, most likely resulting in little measurable impact from co-infection at the population level. Of relevance to the implementation of public health policies, a reduction in RSV circulation, for example, due to vaccination, is therefore unlikely to result in substantial replacement with influenza. This provides evidence against a potential concern for the implementation of an RSV vaccine.

Our model was limited by the need to reduce complexity, and we, therefore, did not allow for waning of immunity, and hence we were unable to include repeat infection of the same virus within a season, or any serological data. Repeat infections of influenza and RSV are likely due to different subtypes. However, as our estimate of interaction is very short-lived, not including these repeat infections are unlikely to have affected our estimates. We were additionally limited by the lack of RSV cases following influenza infections, meaning we had very large confidence on the estimates of the effect of influenza on RSV. We also took a simplistic approach of homogenous population mixing. An alternative explanation of the elevated rate of co-infections is that behavioural aspects may have influenced both the probability of getting RSV and influenza. This could be, for instance, attendance at a crowded event (e.g. religious ceremony) resulting in an increased chance of infection with both viruses. Alternatively, it may just be individuals with higher contacts due to factors such as their employment type. Ideally, we would be able to quantify times of exposure to the viruses, for example, through infections in the household, however, we did not have sufficient data to analyse this in the current study. We also did not look at climatic factors specifically, instead assuming they were captured by our time-varying FOI. This time-varying FOI also allowed us to account for the fact that the influenza season follows the RSV season. We also note that all but one overlapping episode for RSV and influenza were collected during the first season. This is likely due to differing subtypes or age factors, which we were not able to include in our crude analysis of expected dual infections. We attempted to run a sensitivity analysis excluding the first season of data where the majority of co-infections were detected, however this model did not converge, likely due to small numbers.

Overall, the use of a mathematical model combined with a highly detailed longitudinal population-based infection study allowed us to estimate the interaction of RSV and influenza in South Africa. Our findings suggest that concerns for increased influenza circulation as a result of the introduction of RSV prevention strategies may not materialise.

## Data Availability

A summarised version of the data is available at https://github.com/NaomiWaterlow/markov_model

## Funding acknowledgment

The study was funded through a cooperative agreement with the United States Centers for Disease Control and Prevention (CDC) (grant number 1U01IP001048). Testing for RSV was supported by the Bill and Melinda Gates Foundation

(Grant number: OPP1164778). The data analyses were supported by Germany’s Innovation Fund of the Joint Federal Committee (grant no. 01VSF18015).

