## Supplemental Material for "Transient increased risk of influenza infection following RSV infection in South Africa; findings from the PHIRST study, South Africa, 2016-2018"

**Contents:**

- PCR methods
- Risk ratio calculations
- State definitions
- Transition between states
- Misclassification matrix
- Sensitivity Analyses

1. **PCR methods**

Nucleic acids were extracted using the Roche MagNA Pure 96 instrument (Roche, Mannheim, Germany) according to the manufacturer’s instructions. Nasopharyngeal samples were tested for influenza A and B viruses, and RSV by real-time reverse transcription polymerase chain reaction (rRTPCR) using the FTD Flu/RSV detection assay (Fast Track Diagnostics, Luxembourg). Influenza A-positive samples were subtyped using the Centers for Disease Control and Prevention (CDC) influenza A (H1/H3/H1pdm09) subtyping kit and influenza B lineage was determined using the CDC B/Yamagata-B/Victoria lineage typing kit (available through Influenza Reagent Resource Program; [www.influenzareagentresource.org](http://www.influenzareagentresource.org)).

2. **Risk ratio calculations**

Risk ratios and confidence intervals were calculated using the following formulas:

Risk ratio = (a/(a+b)) / (c/(c+d))

Lower 95% = exp(log(risk ratio) + 1.96* sqrt(((b/a)/(b+a)) + ((d/c)/(d+c))))

Upper 95% = exp(log(risk ratio) - 1.96* sqrt(((b/a)/(b+a)) + ((d/c)/(d+c))))

Where a, b, c, and d are the contents of a two-by-two table:

|  | **With outcome (positive sample for second virus)** | **Without outcome (no positive sample for second virus)** |
| --- | --- | --- |
| **Exposed group (positive sample for primary virus)** | a | b |
| **Unexposed group (negative sample for primary virus)** | c | d |

3. **State definitions**

States are defined as follows:

SS - Susceptible to RSV and Susceptible to influenza

SPR - Susceptible to RSV and Protected or Recovered from influenza

PRS - Protected or Recovered from RSV and Susceptible for influenza

RR - Recovered from RSV and Recovered from influenza

SI - Susceptible to RSV and Infected with influenza

IS - Infected with RSV and Susceptible to influenza

IPR - Infected with RSV and Protected or Recovered from influenza

PRI - Protected or Recovered from RSV and infected with influenza

II - Infected with RSV and influenza

RR - Recovered from RSV and influenza

Upon enrollment in the study each individual is assumed to be in State SS. The first infection time of each individual for each virus are denoted $\tau_{i,RSV}$ and $\tau_{i,Flu}$ respectively and the last infection time of each virus are $\eta_{i,RSV}$ and $\eta_{i,Flu}$, and the following rules apply, where $t$ denotes the swab date

$t < \tau_{i,Flu} \& t <\tau_{i,RSV}$, state = SS

$t > \tau_{i,Flu} \& t <\tau_{i,RSV}$, state = SPR

$t < \tau_{i,Flu} \& t >\tau_{i,RSV}$, state = PRS

$t > \tau_{i,Flu} \& t >\tau_{i,RSV}$, state = RR

$t == \tau_{i,Flu} \& t <\tau_{i,RSV}$, state = SI

$t < \tau_{i,Flu} \& t ==\tau_{i,RSV}$, state = IS

$t > \tau_{i,Flu} \& t ==\tau_{i,RSV}$, state = IPR

$t == \tau_{i,Flu} \& t >\tau_{i,RSV}$, state = PRI

$t == \tau_{i,Flu} \& t ==\tau_{i,RSV}$, state = II

$t > \eta_{i,Flu} \& t >\eta_{i,RSV}$, state = RR

And that IPR and PRI are censored states, as we cannot distinguish between IP and IR and or between PI and RI in the order of events.

4. **Transitions between states**

Transition between states is governed by the Q matrix below, where the order of the states is: SS, IS, PS, RS, SI, II, PRI, SP, IPR, SR, RR.
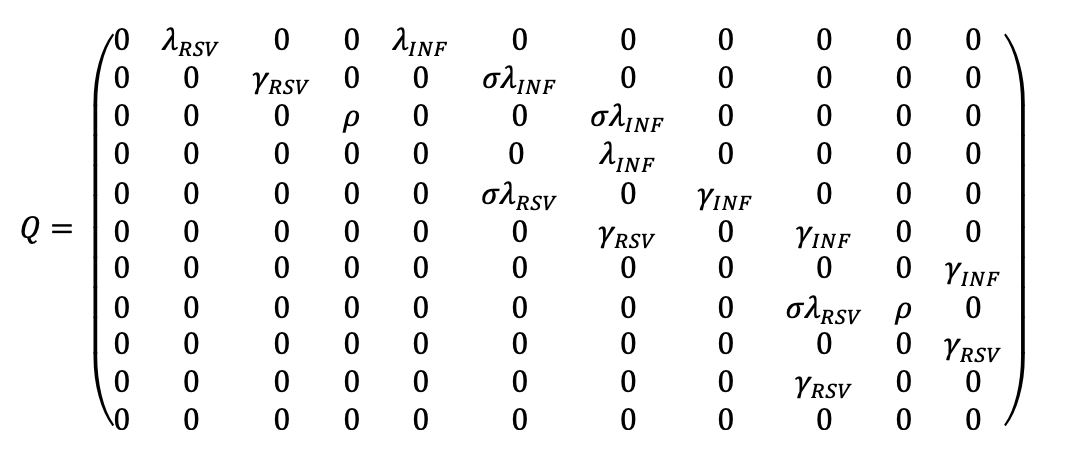

5. **Misclassification matrix**

We allow misclassification to occur for negative swabs, but take all positive swabs as true positives. This results in the following misclassification (E) matrix, where $\epsilon$ denotes the probability of each state (row) being misclassified as a different state (column). The $\delta$ for each state indicates 1 - the sum of the misclassification rates in that column.

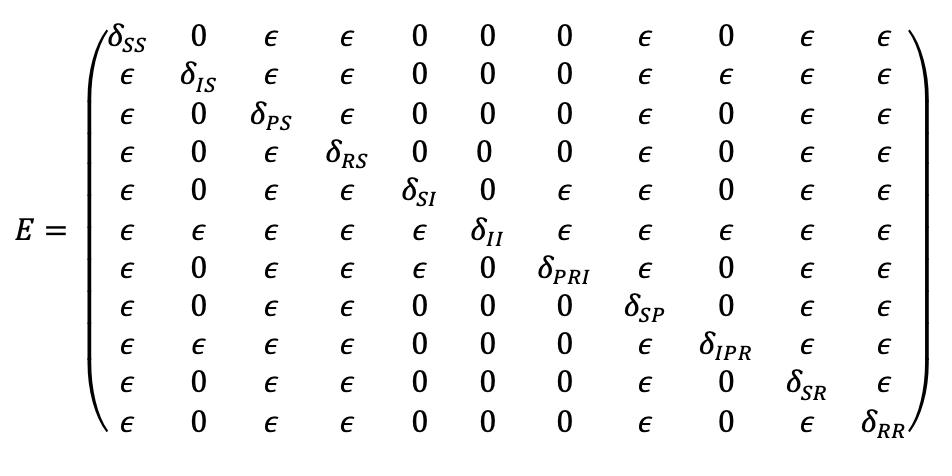

6. **Model Fit**

Table 1 shows the fitted model parameters and figure S1/2 shows the model estimates against observed input data.

*Table 1: Estimated parameter values from the main model.*

| **Model** | **Interaction strength influenza -> rsv** | **Interaction strength rsv -> influenza** | **Initial state occupancy probability for state SS** | **False negative test misclassification probability** | **-2 log likelihood** |
| --- | --- | --- | --- | --- | --- |
| **7 days** | 0.93 (0.2 - 4.3) | 2.13 (0.97 - 4.69) | 0.998 (0.876-0.998) | 0.007481 (0.00724 - 0.00773) | 56717.79 |
| **7 days (Replicate)** | 1.38 (0.45 - 4.26) | 2.03 (0.90 - 4.56) | 0.998 (0.586-0.998) | 0.00748 (0.00724 - 0.00773) | 56719.61 |

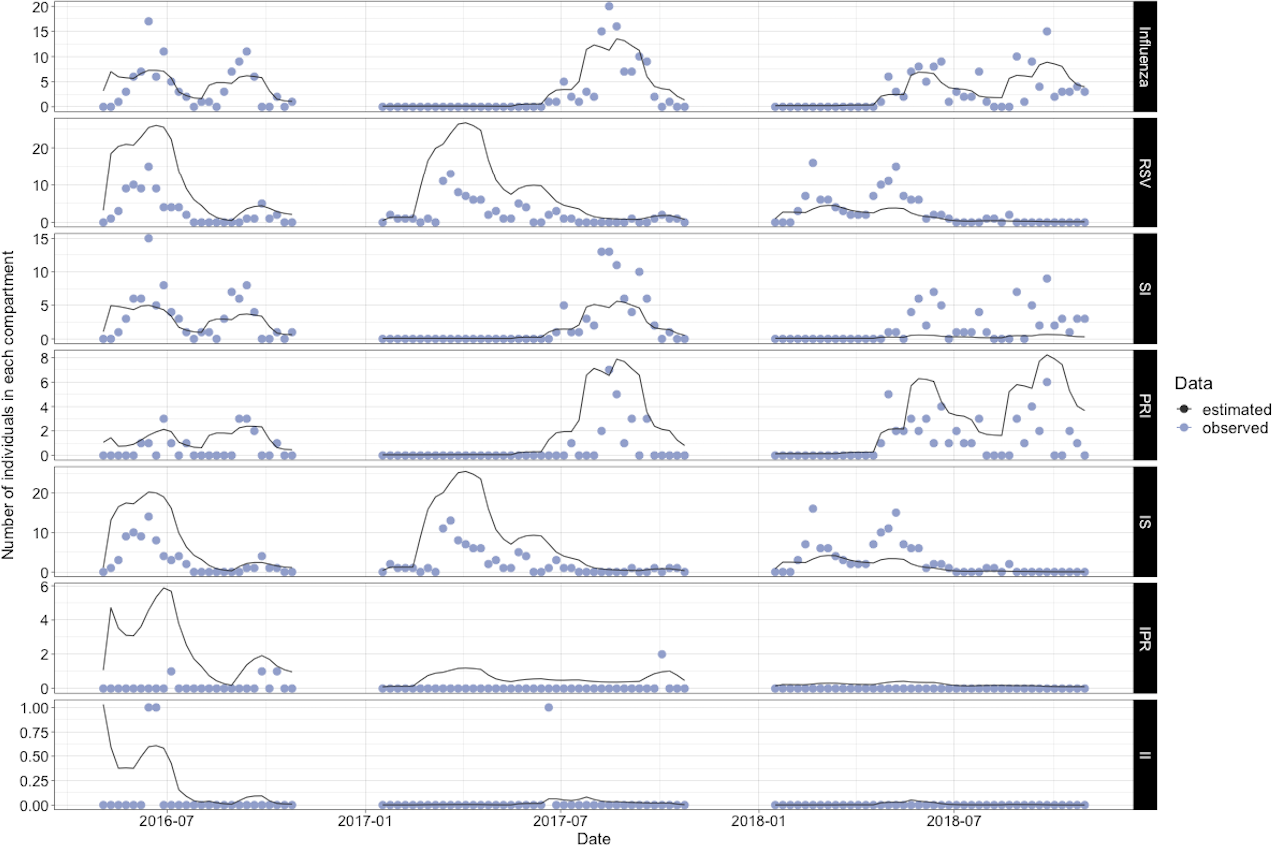

*Figure S1: Model Fit. Count of individuals in each infected compartment for data (blue) against model forecasts (black). The first two rows indicate the overall number of influenza and RSV infections respectively, followed by the compartment-specific counts. The compartment acronym are: SI = Susceptible RSV, Infected influenza, PRI = Protected or Recovered RSV, Infected influenza, IS = Infected RSV, Susceptible influenza, IPR = Infected RSV, Protected or Recovered influenza and II = Infected with both influenza and RSV. For non-infected compartments and sensitivity analyses see supplement.*

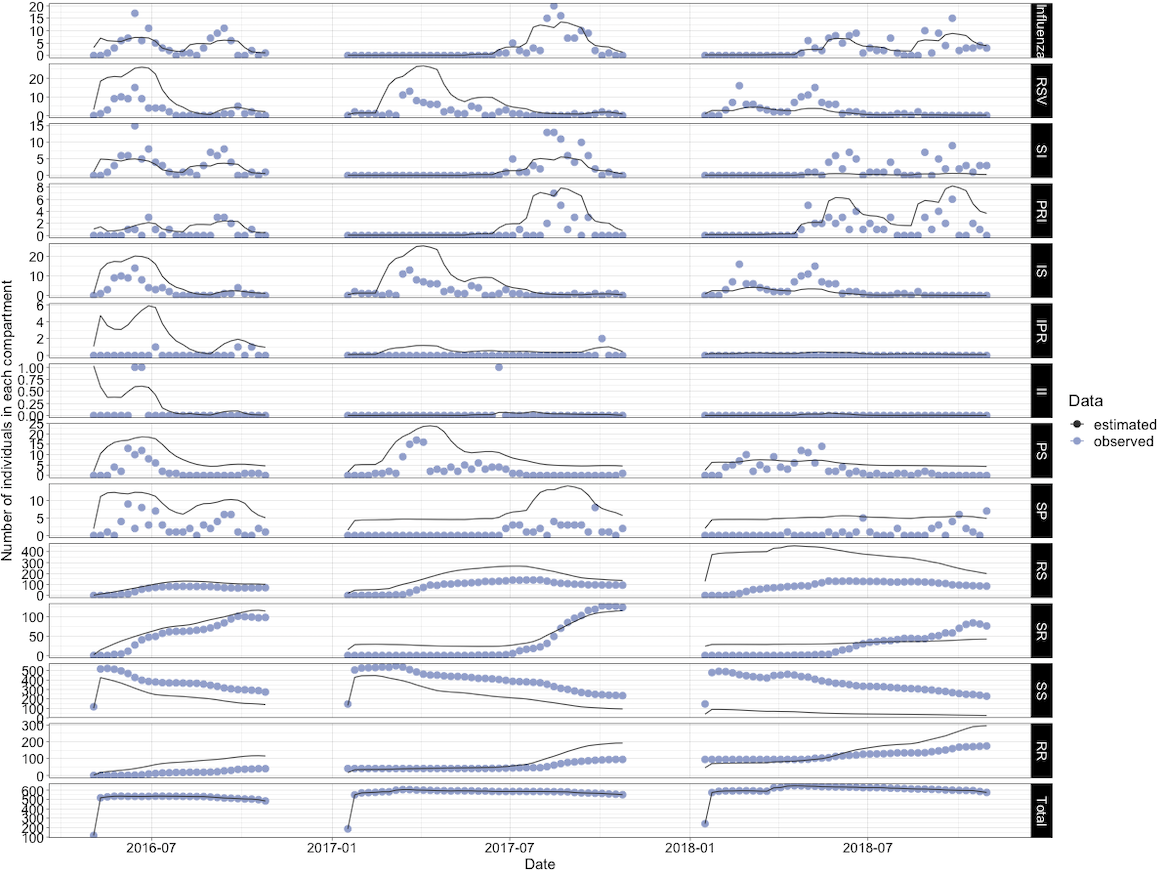

*Figure S2: Model Fit. Count of individuals in each infected compartment for data (blue) against model forecasts (black). The first two rows indicate the overall number of influenza and RSV infections respectively, followed by the compartment-specific counts. The compartment acronym are: SI = Susceptible RSV, Infected influenza, PRI = Protected or Recovered RSV, Infected influenza, IS = Infected RSV, Susceptible influenza, IPR = Infected RSV, Protected or Recovered influenza, II = Infected with both influenza and RSV, PS = Protected RSV, Susceptible influenza, SP, Susceptible RSV, Protected influenza, RS = Recovered RSV, Susceptible influenza, SR = Susceptible RSV, Recovered influenza, SS = Susceptible RSV and influenza, RR = Recovered RSV and influenza.*

7. **Sensitivity Analyses**

Table S1 shows the model results for the default model (7 days interaction) and the sensitivity analyses, where the duration of interaction $\rho$ was fixed at 14, 21 and 28 days respectively. The relevant Force of Infections and age group hazard rates are shown in Figures S3 - S5.

| **Model** | **Interaction strength influenza -> rsv** | **Interaction strength rsv -> influenza** | **Initial state occupancy probability for state SS** | **False negative test misclassification probability** | **-2 log likelihood** |
| --- | --- | --- | --- | --- | --- |
| **14 days** | 0.96 (0.3 - 3.04) | 1.76 (0.88 - 3.54) | 0.997 (0.114 - 0.998) | 0.00748 (0.00724 - 0.00773) | 56718.35 |
| **14 days (Replicate)** | 0.88 (0.26 - 2.93) | 1.98 (1.04 - 3.77) | 0.998 (0.463 - 0.998) | 0.00748 (0.00724 - 0.007732 | 56721.04 |
| **21 days** | 0.93 (0.34 - 2.55) | 1.72 (0.92 - 3.19) | 0.977 (0.876 - 0.998) | 0.00748 (0.00723 - 0.00773) | 56718.11 |
| **21 days (Replicate)** | 0.69 (0.2 - 2.42) | 1.76 (0.96 - 3.24) | 0.998 (0.315 - 0.998) | 0.00748 (0.00724 - 0.00773) | 56720.04 |
| **28 days** | 0.89 (0.35 - 2.26) | 1.78 (1.02 - 3.09) | 0.997 (0.145 - 0.998) | 0.00748 (0.00724 - 0.00773) | 56717.52 |
| **28 days (Replicate)** | 0.79 (0.29 - 2.15) | 1.77 (1.01 - 3.08) | 0.998 (0.547 - 0.998) | 0.00748 (0.00724 - 0.00773) | 56720.18 |

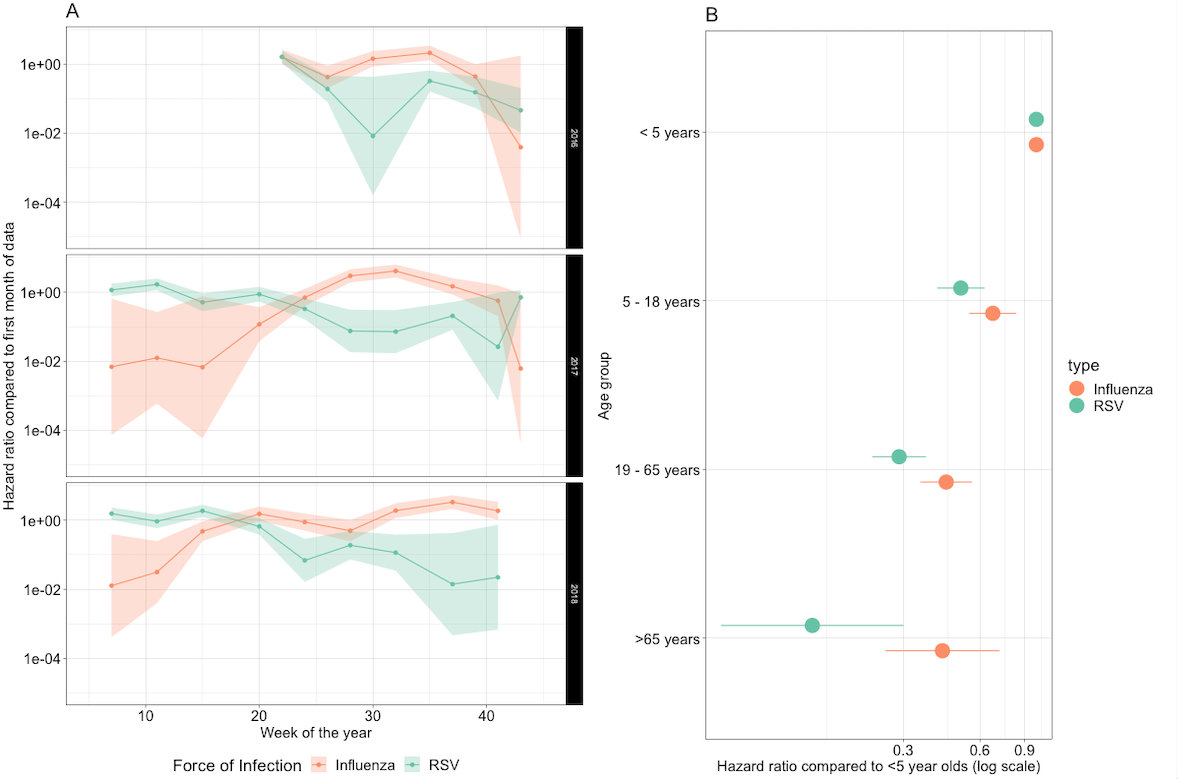

Figure S3: *A) Model output time dependent Force of infection for influenza and RSV (interaction duration = 14 days). Lines indicate the mean value and the shaded ribbons the 95% CI. B) Age group Hazard ratios for susceptibility of infection, with age group <5 years old as the base age group.*

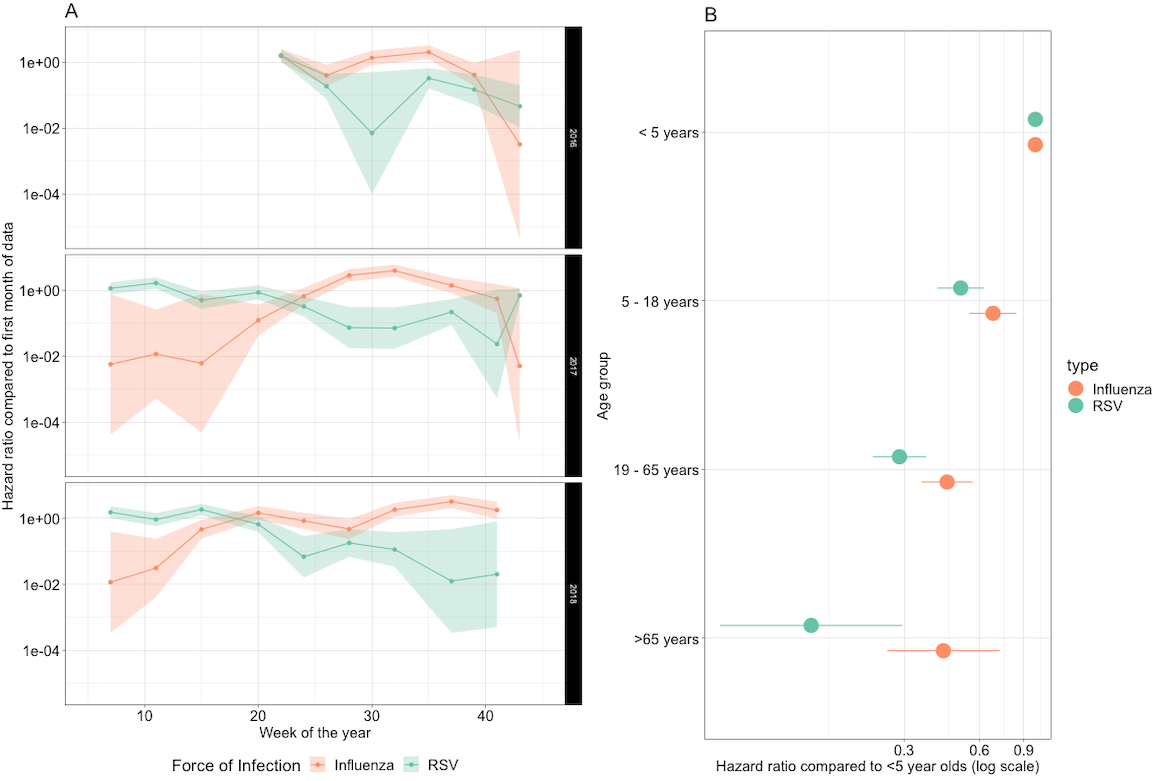

Figure S4: *A) Force infection over time for influenza and RSV (interaction duration = 21 days). Lines indicate the mean value and the shaded ribbons the 95% CI. B) Age group Hazard ratios, with age group <5 years old as the base age group.*

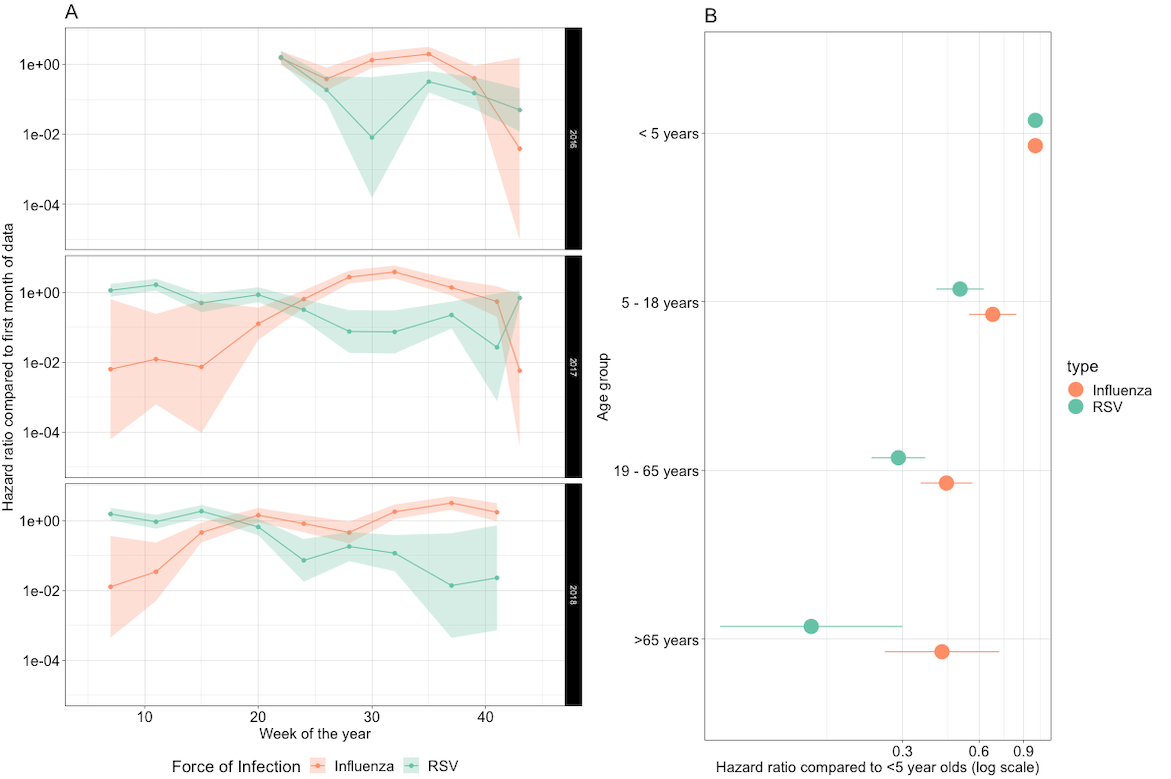
Figure S5: *A) Force infection over time for influenza and RSV (interaction duration = 28 days). Lines indicate the mean value and the shaded ribbons the 95% CI. B) Age group Hazard ratios, with age group <5 years old as the base age group.*
